# Towards chronomic medicine: Enrichment and linkage of chronotype markers with rare disease associated genes

**DOI:** 10.1101/2020.08.17.20176420

**Authors:** Basimah T Malik, Timothy J Hearn

**Affiliations:** Academic Department of Medical Genetics, University of Cambridge; Research Department of Cell and Developmental Biology, University College London

**Keywords:** Chronotype, linkage, enrichment, Long QT Syndrome, OMIM, HPO

## Abstract

The role of the circadian clock is becoming apparent in many aspects of human health and disease. Large scale GWAS studies have delivered high numbers of genetic markers for chronotype, which can be used to find links to Mendelian disorders. We used the variants in the 1,000 genomes study to estimate linkage disequilibrium for these chronotype markers. We analysed genes in high linkage disequilibrium with the chronotype markers for enrichment of disease-causing genes, and looked for enrichment of common HPO terms in the wider disease associated genes. We identified that two cardiovascular disorders, cardiomyopathy, and the inherited cardiac arrhythmia Long QT Syndrome, along with the immune system disorder complement component 2 deficiency were significantly enriched MIM diseases. In contrast the most common HPO terms were developmental and neurological terms. This analysis provides a starting point for identifying the circadian contribution to disease outside of the core circadian clock genes, by providing candidate conditions and loci for identifying circadian modifier variants.

## Introduction

Circadian rhythms are biological rhythms that are endogenous in nature and persist in constant conditions, driven internally by an organism’s circadian clock. Humans contain an intrinsic circadian clock that co-ordinates physiology with the external environment, and is increasingly seen as having a pervasive role in human health and disease (Roenneberg and Merrow 2016). The master circadian pacemaker is located in the suprachiasmatic nucleus (SCN) of the hypothalamus and synchronizes peripheral oscillators in other tissues through regulating rhythmic endocrine hormone release. However, the oscillators within individual tissues and organs drive their own distinct expression patterns of rhythmic gene expression (Ruben et al., 2018).

Due to the focus on neuroendocrine systems in regulating the circadian system much circadian research in humans has been performed on sleep-wake schedules. The concept of chronotype has been developed to describe an individual’s circadian phase based on the timing and duration of their sleep wake cycle during the 24 hour period (Roenneberg et al., 2007). An increasing number of clinical associations are being found with chronotype, such as body mass index (Maukonen et al., 2016). Over the past decades an increasing number of genetic markers have been identified that are enriched in or associated with individuals with early or late chronotypes. Recent GWAS studies have led to an extended list of 351 chronotype markers based on the large UK biobank and 23 and me datasets (Jones et al., 2019).

Logically much research into human circadian clocks has focused on neuroendocrinology, due to the location of the master pacemaker in the SCN, and the utility of using circadian timing information based on timing of melatonin release in chrono therapeutics (Roenneberg and Merrow 2016). The relevance of the circadian clock with rare disease is a comparatively unexplored area. Here we begin to probe the beginnings of identifying clinically relevant variants in the circadian system, by looking at the clinical relevance of the chronotype marker variants using a genomic medicine approach.

Linkage disequilibrium (LD) is a useful tool for identifying genes linked to intergenic markers, and for identifying variants likely to be inherited together. The linkage disequilibrium tools from the 1,000 genomes programme (Auton et al., 2015) allow this to be estimated for the many sub populations of this study, and therefore predict the genes where variants are likely to be linked to chronotype markers in these populations.

The basic complement of genomic medicine resources, such as OMIM, and ClinVar are useful resources for identifying the clinical relevance of genes and variants, and can from the basis for identifying enrichment of genetic disorders. Newer concepts such as the Human Phenotype Ontology allow a more nuanced analysis to be performed by identifying common phenotypes between diseases

Here we identify the genetic conditions and phenotypes that are most enriched in genes associated with variants linked to chronotype markers. This is our first step in bringing chronobiology and genomic medicine together in ‘chronomic’ medicine.

## Methods

### Identification of 1000 genomes variants in linkage disequilibrium with chronotype markers

The Ensembl Linkage Disequilibrium calculator was used to estimate linkage disequilibrium (LD) in the 1,000 genomes (1KG) population (https://www.ensembl.org/Homo_sapiens/Tools/LD). The calculator was set to give ‘LD for a given variant and defined window size’, which was set as 20000. All 1KG populations were selected. The rsIDs for the 351 chronotype markers previously identified (Jones et al., 2019) were used as input, except rs67169439, which was not found in 1,000 genomes. The results for all populations were exported and sorted in order of *r*^2^ values and variants with an *r*^2^ value greater than 0.95 and a D’ value greater than 0.95 were retained for further analysis. Only after this were all duplicate rsIDs removed, to ensure that all wanted variants remained in the list and were not deleted before the *r*^2^ filtering due to different LD in the 1000 genomes sub populations

### Clinical Significance, Gene Stable ID and PubMedID of variants in linkage disequilibrium with the chronotype markers

The unique rsIDs for variants in LD with the chronotype markers from the previous analysis were entered as filters into the Ensembl Biomart https://www.ensembl.org/biomart/martview/ (Yates et al., 2019) ‘Human Short Variants (SNPs and indels excluding flagged variants) (GRCh38.p13) dataset ‘Ensembl Variation 100’ release. The ‘Variant name’, ‘Variant source’, ‘Clinical significance’, ‘PubMed ID’, ‘Gene name’ and ‘Gene Stable ID’ attributes were extracted.

### Chronotype-linked Associated Genetic Disorders and enriched genes

By using the Gene Stable IDs from the results from the previous experiment a list of genes was compiled. These were used as filters for the Ensembl biomart https://www.ensembl.org/biomart/martview/ (Yates et al., 2019) ‘Ensembl Genes 100’ release of the ‘Human genes (GRCh38.p13)’ dataset. The attributes were set to output ‘Gene Stable ID’, ‘MIM morbid description’ and ‘MIM morbid accession’. These were quantified to identify multiple occurrences of OMIM entries.

The list of gene names obtained from Ensembl were entered into Enrichr https://amp.pharm.mssm.edu/Enrichr/ (Kuleshov et al., 2016) (Chen et al., 2013). Using the Diseases/Drugs tab the OMIM disease table was exported to get the Fisher’s p value and odds ratio. These are quoted in the text as p-value and OR. As well as this, the related clustergram was also downloaded for OMIM disease, ordered by enrichment sum.

Finally, the corresponding gene names were added to the MIM morbid table. The genes were then compared to the enriched genes in the table exported from Enrichr, any genes that were in the table (and so were enriched) were removed from the list, leaving a list of genes that were not enriched.

### Quantifying the HPO terms for chronotype linked genes

The previous “MIM morbid accession” numbers were inputted into the HPO term site https://hpo.jax.org/app/, (Köhler et al., 2019) and then from the disease results all of the HPO term associations were downloaded. For any ‘MIM morbid accession’ numbers that appeared more than once (i.e. 2 different genes gave the same MIM number), the HPO results for that MIM number were duplicated to match the number of MIM numbers. The HPO terms were copied and compiled into a list. The number of times each term appeared in the list was quantified.

## Results

### Identification of variants in high Linkage Disequilibrium with chronotype markers

To identify the association of chronotype markers with genetic disease we performed linkage disequilibrium analysis using the 1,000 genomes project data. Linkage disequilibrium (LD) was calculated for the 350 of the 351 chronotype markers identified in previous GWAS studies (Jones et al., 2019) that are present in the 1,000 genomes project populations (Supplemental table 1). A total of 13,838 variants were found in LD with *r*^2^ and D’ above 0.95. The variants in high LD were assessed for clinical significance (Table 1). Of the 13,838 variants, 4318 had citations in PubMed and 44 variants had at least one clinical assessment. Only one variant, rs1815739 had an indication of pathogenicity with a mixed benign,pathogenic report. The high LD variants were then filtered for gene IDs to identify the genes in LD with the chronotype markers in the different populations of the 1,000 genomes project. 11678 variants were in protein coding genes, which corresponded to 433 unique genes.

**Table 1.**
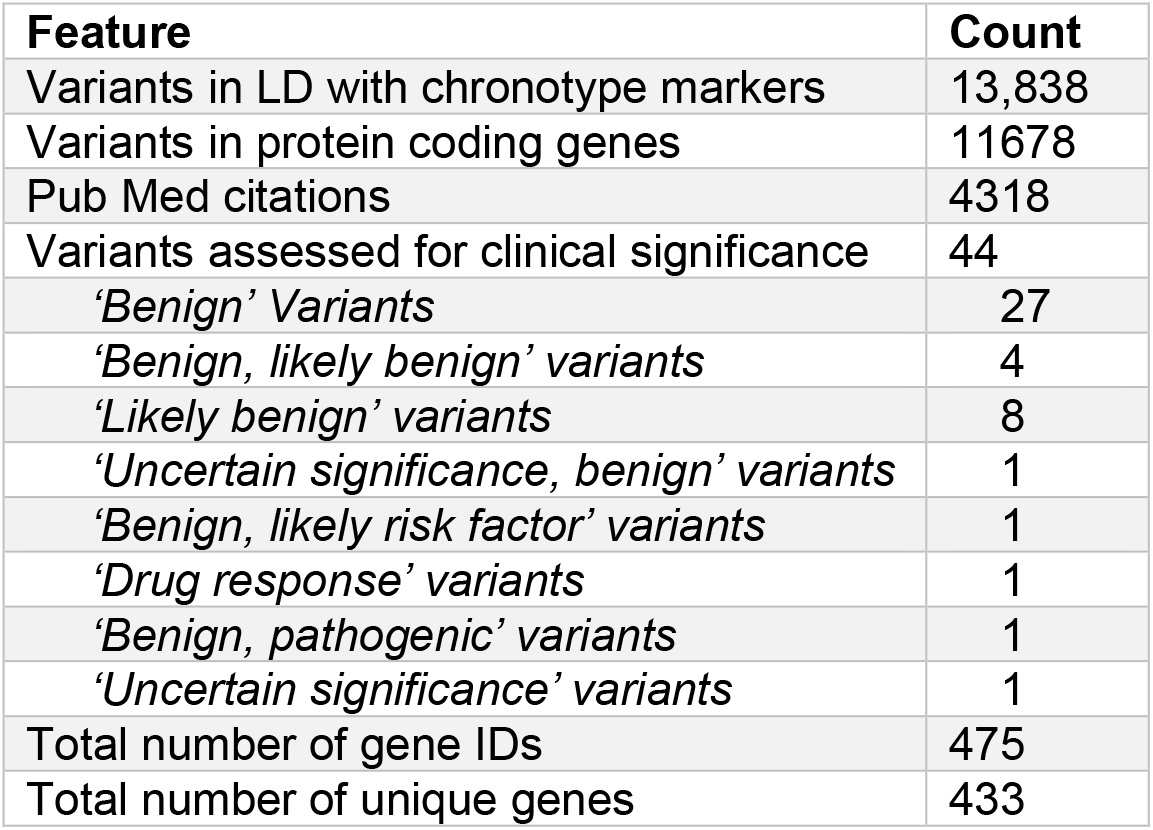
Summary of the variants in linkage disequilibrium with chronotype markers in the 1,000 genomes project. The number of variants in LD with chronotype markers calculated from the 1KG populations were assessed for PubMed citations, clinical significance in ClinVar and the genes that those variants are associated with.

### Enrichment for diseases associated with chronotype-linked genes

The MIM terms associated with these chronotype-linked genes were extracted (Supplemental table 2) and the 187 MIM morbidity numbers quantified (Table 2). Fourteen diseases appeared multiple times associated with the chronotype-linked genes (Supplemental table 2). The chronotype-linked genes were analysed for enrichment for specific diseases using the MIM terms. The most enriched disease terms are shown with the associated genes in figure 1. The significantly enriched terms were hypertrophic cardiomyopathy (P value: 0.005 OR: 8.15), Long QT Syndrome (P value: 0.026 OR: 7.69) and complement component 2 deficiency (P value: 0.031 OR: 7.11). 91 genes were not associated with an enriched disease.

**Table 2.**
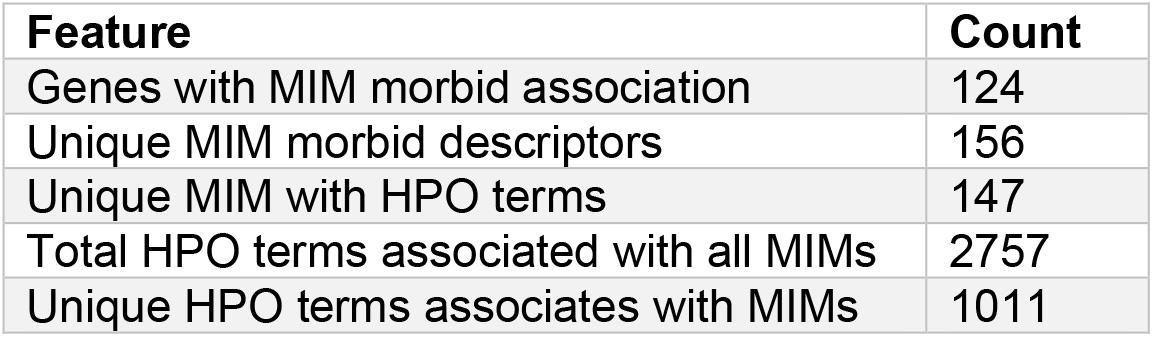
MIM morbid and HPO term enrichment in chronotype-linked genes. The genes associated with chronotype linked variants were assessed for Mendelian inheritance in man (MIM) morbidity terms. These terms were used to generate human phenotype ontology (HPO) terms associated with the genes.

**Figure 1.**
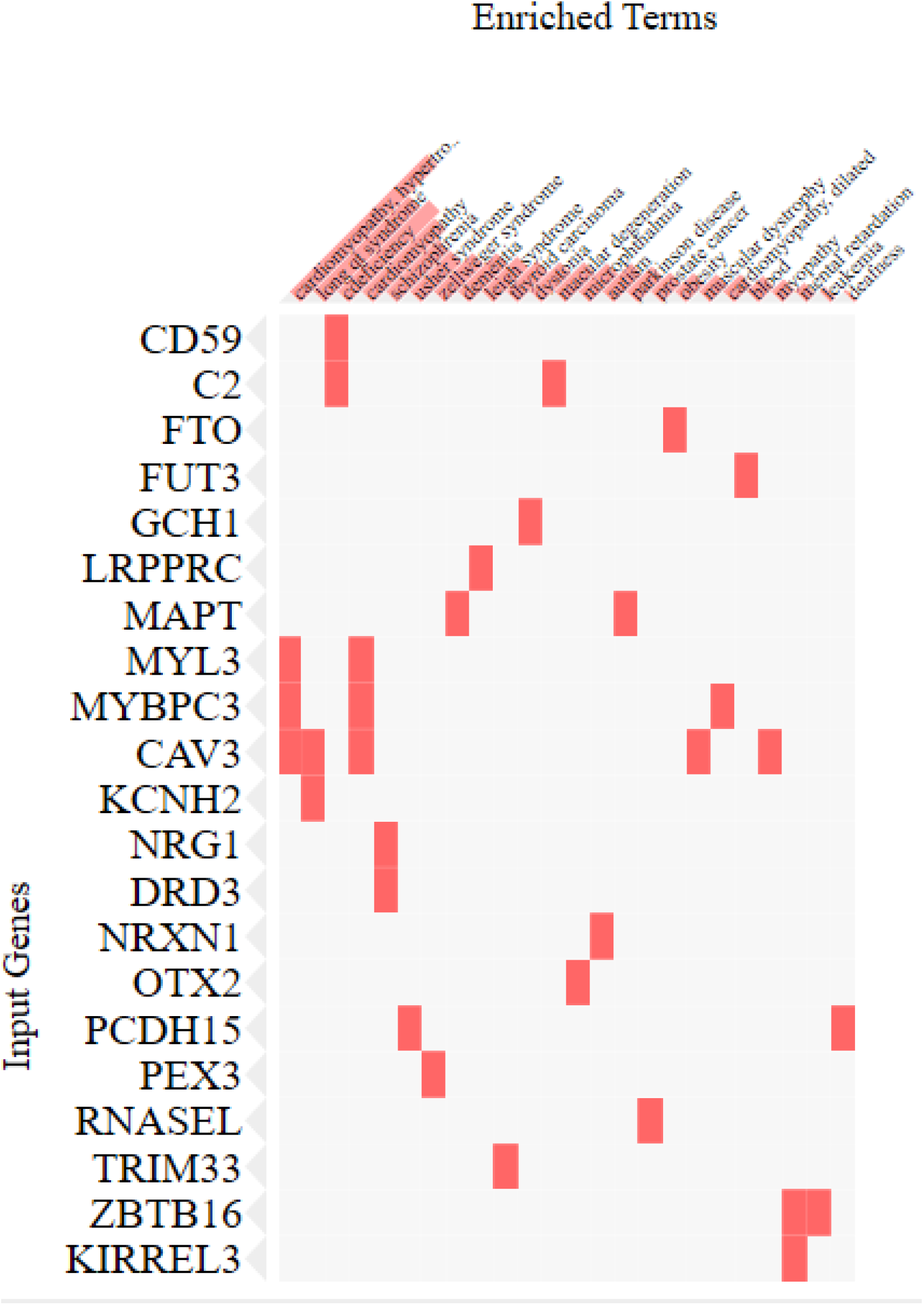
Enrichment of MIM terms with associated genes in the chronotype-linked genes. Enrichr was used to calculate enrichment using Fisher’s exact test. The MIM terms on the top x axis are organized by order of enrichment p value (Supplemental Table 2). Gene names underlying enrichment are shown in the clustergram.

The use of MIM terms for enrichment restricts enrichment analysis to particular disease sub-groups. An alternative method is to use the HPO terms associated with the MIM terms to quantify similar phenotypes between diseases.

### Enrichment of HPO terms in diseases associated chronotype-linked genes

The HPO terms associated with the MIM terms were extracted and quantified. There were 1011 HPO terms associated with 147 unique MIMs. Of these 560 terms were unique. The highest occurring terms were to do with the genetic inheritance 101 autosomal dominant and 83 autosomal recessive terms. The highest occurring disease associated terms were terms associated with neurological and developmental disorders (Table 3). The full list is shown in Supplemental Table 3.

**Table 3.** Highest occurring HPO terms shared by MIM terms found for chronotype-linked genes. Count of HPO terms associated with MIM terms for chronotype-linked genes.

| HPO term | Count |
| --- | --- |
| Global developmental delay | 44 |
| Generalized hypotonia | 41 |
| Seizure | 35 |
| Intellectual disability | 31 |
| Dysarthria | 22 |
| Strabismus | 21 |
| Short stature | 20 |
| Muscular hypotonia | 19 |
| Delayed speech and language development | 18 |
| Microcephaly | 17 |

### Manual curation of chronotype variants associated with enriched MIM terms

We have identified the disease association with variants in LD with the previously identified chronotype markers, but the clinical significance of the chronotype markers themselves is also of interest. The 351 chronotype variants were searched for clinical significance using Biomart, which yielded only two entries; 1 likely benign and 1 risk factor variants. However, clinical significance is not always completely annotated in ClinVar, and a manual search for the variants associated with the enriched MIM terms was performed. Of these rs2072413 (in *KCNH2*) was identified from two publications as modulating the QT interval in enrichment studies (Corponi et al., 2019; Arking et al., 2014).

## Discussion

In this analysis we identified the genes associated with variants under linkage disequilibrium with the chronotype markers discovered in recent GWAS (Jones et al. 2019). We found that these genes were enriched for two cardiovascular diseases and one immune deficiency disorder. The cardiovascular disorders hypertrophic cardiomyopathy and Long QT syndrome were found to be the most enriched MIM terms. Both cardiomyopathy and Long QT syndrome are good candidates for diseases where there might be a circadian contribution to pathophysiology. Both the heart rate and the QT interval are known to exhibit diel oscillation patterns (Black et al., 2019). Genetic modifiers of the nocturnal increase in QT interval have not been reported, however, one of the chronotype markers behind the enrichment in Long QT syndrome associated genes in this analysis, rs2072413, has been found to modulate QT prolongation previously (Corponi et al., 2019; Arking et al., 2014), indicated a potential functional link to nocturnal QT lengthening that should be considered in future studies. Different Long QT syndromes have different timing of cardiac events. Our enriched MIM term corresponds to genes associated with Long QT 9 (CAV3) and Long QT 2 (ALG10B, KCNH2). Long QT syndrome 2 events are known to be associated with the early part of the day (Black et al., 2019), potentially indicating a role for chronotype in this disease.

The enrichment in complement component 2 deficiency associated genes is intriguing as increased inflammation to communicable infections such as influenza have been shown to be correlated with the evening (Punder et al 2019). The circadian clock plays a critical role in human immune system homeostasis, with immune responses mediated time dependently. The role of the circadian clock in governing immune responses to communicable diseases is rapidly becoming established (Mazzoccoli et al., 2020; Zhuang et al., 2017), with circadian rhythm modulation characterised for both influenza related lung inflammation and survival due to time of original infection (Sengupta et al., 2019). The severity of acute influenza infections is governed by the circadian clock as disruption leads to increased influenza viral replication and transmission (Edgar et al.,2016). Better survival of influenza infection is associated with the time dependent gating of the quantity of NK and NKT cells and inflammatory monocytes in the lungs (Sengupta et al., 2019). Thus the disorders identified through our MIM enrichment are examples of diseases where diurnal and circadian regulation of the underlying pathophysiology is already well known.

Our HPO analysis identifies the restriction of using MIM terms alone for disease enrichment, and identifies a different set of disease features that could be considered for identifying circadian contribution to rare disease. Diseases were split approximately 50% between recessive and dominant disorders. The most common feature were aspects associated with developmental delay. The inclusion of short stature as a marker is of interest, as studies have utilized height measurements to assess and demonstrate that chronotype affects BMI (Maukonen et al., 2016). Additionally, both height and chronotype appear to be associated with season of birth (Didikoglu et al., 2020), indicating some level of photoperiodic influence on development.

One chronotype-linked variant, rs1815739, was shown in the results to be ‘benign, pathogenic’. This variant belongs to ACTN3 (Actinin alpha 3), a gene which is mostly expressed in the Z line of sarcomeres, where actin is bound onto in skeletal muscle. The ClinVar (Landrum et al., 2018) entry for this variants had opposing ‘interpretations’, some evidence points towards rs1815739 being benign and having no effect on an individual, in fact (North et al., 1999) shows ACTN3 to be “functionally redundant”. Other research suggests the variant is less prevalent amongst athletes and so must be disadvantageous (Roth et al., 2008). The variant has also been associated with myopathies, for example Duchenne Muscular Dystrophy (Houweling et al., 2018).

Together, these results indicate genetic diseases that should be prioritized as promising candidates for future analysis of the circadian contribution or modulation of disease status. The variants with associations highlighted here represent a starting point for this future analysis.

## Data Availability

All data used in analyses is presented in supplemental tables 1-3

## Acronyms and abbreviations

OMIM: Online Mendelian Inheritance in Man
HPO: Human Phenotype Ontology
ICA: Inherited cardiac arrhythmia
GWAS: Genome Wide Association Study LD – Linkage Disequilibrium
1KG: 1,000 Genomes Project

## Acknowledgements

BTM was generously supported by a Genetics Society 2020 studentship awarded to TJH

---

Supplemental Table 1 – List of variants in high LD with chronotype markers and associated genes

Supplemental Table 2 – List of MIM terms associated with chronotype-linked genes

Supplemental Table 3 – List of HPO quantified terms.

